# *TP53* mutation screening for patients at risk of myeloid malignancy

**DOI:** 10.1101/2024.02.06.24302401

**Authors:** Devdeep Mukherjee, Rialnat A. Lawal, Courtney D. Fitzhugh, Christopher S. Hourigan, Laura W. Dillon

## Abstract

There is increasing recognition of the risk of developing therapy-related myeloid malignancy, including after cellular therapy. While retrospective studies have implicated pre-existing *TP53* mutated hematopoietic clones as a common causative mechanism, no prospective screening to identify those patients at greatest risk is currently possible. We demonstrate that ultradeep DNA-sequencing prior to therapy may be used for discovery of *TP53* mutations that are subsequently associated with malignancy.

## MAIN TEXT

Therapy-related myeloid malignancy (tMN) is a devastating, often fatal, complication of otherwise curative therapy. It accounts for approximately 10% of all myelodysplastic syndrome and acute myeloid leukemia (AML) cases and is thought to be commonly induced by positive selection of pre-existing clones^1^. Chromosomal monosomy, a complex karyotype, and mutations in the *TP53* gene are the most frequent genetic aberrations in tMN^2,3^. Latency ranges from several months to years^4^.

Sickle cell disease (SCD) is a group of inherited red blood cell disorders associated with significant morbidity and early mortality. Hematopoietic cell transplantation (HCT) is a curative option for SCD but is associated with a risk of tMN, particularly following graft rejection^5^. Pretransplant conditioning regimens may select for chemo-resistant pre-malignant clones existing below the detectable range of commonly used clinical methods. We have shown previously that pre-existing clones containing *TP53* mutations in SCD patients undergoing HCT can progress into tMN months to years after transplant^6^. Given the increasing use of cellular therapy to treat a variety of diseases, including HCT for SCD, there is an immediate need to develop screening methods for discovery of pre-existing low level hematopoietic clones with the potential for evolution into tMN^7,8^.

To fulfill this unmet need, we developed an ultra-deep DNA sequencing panel targeting the entire coding sequence of the *TP53* gene (Figure 1A). This methodology utilizes single-strand unique molecular indices for error-corrected consensus calling, which in combination with background error modeling, allows for confident, highly specific, discovery of pathogenic variants far below the normal limit of detection for standard next generation sequencing (NGS) assays. While known variants can often be detected with conventional NGS, the false positive error rate precludes discovery of previously unknown mutations present at low variant allele fractions (VAF). Assay performance was assessed by serial dilutions of known *TP53*-mutated positive control samples into healthy human donor blood DNA with anticipated VAFs ranging from 0.003% to 1%. Focusing on the most frequently mutated regions of *TP53*, including 62 known hotspots (Figure 1B, Supplementary Table 1) ^9,10, 11,12,13^, we found that the median error corrected sequencing depth in these regions was 14,512 (95% CI: 14,769 – 15,430) (Figure 1C) with a median theoretical *de novo* discovery limit of 0.058% (Figure 1D). Experimentally, the *TP53* NGS assay could detect 7 *TP53* mutations, including 4 missense (c.536A>G, c.537T>G, c.725G>C, c.818G>A), 2 splicing (c.375+5G>A, c.993+1G>T) and 1 frameshift (c.370del) variant (Supplementary Table 2 with 100% specificity, down to a lower limit of discovery limit of 0.05 – 0.12% (Figure 1E). Variant dilutions were orthogonally quantified using digital droplet PCR (ddPCR) and showed a high correlation with NGS VAF (*r* = 0.958, *p < 0*.*0001*) (Figure 1E).

**Figure 1.**
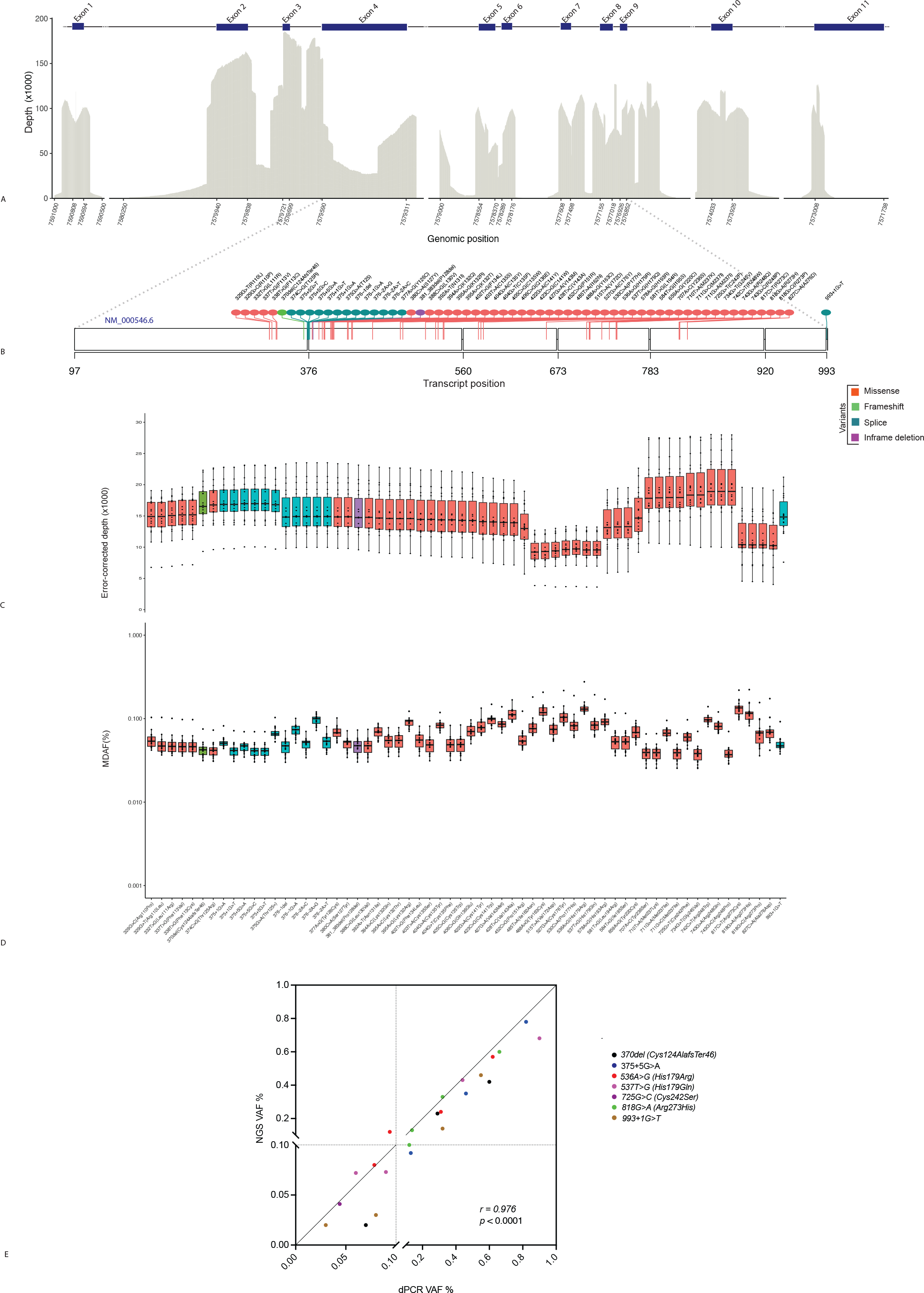
*TP53*-specific NGS. Mean sequencing depth (A) in regions of common TP53 mutational hotspots in AML and t-MN (B). Error-corrected depth (C) and corresponding minimal detectable allele fraction (% MDAF) (D) at each hotspot. Serially dilution of positive control samples with known *TP53* variants positive into healthy donor blood DNA. A total of 500ng DNA was used for both NGS and droplet digital PCR (ddPCR) *r*, Pearson correlation coefficient (E).

Next, we investigated the presence of pre-HCT *TP53* variants in a cohort of 19 SCD patients who underwent non-myeloablative peripheral blood (PB) HCT at the National Institutes of Health between June 2010 and October 2020. HLA-matched sibling HCT included alemtuzumab, 300cGy total body irradiation (TBI), and sirolimus with or without pentostatin and oral cyclophosphamide (PC) preconditioning (Table 1). Haploidentical HCT included alemtuzumab, 400cGy TBI, and sirolimus with or without post-transplant cyclophosphamide (up to 100mg/kg) and PC preconditioning. Eleven of 19 patients experienced graft failure (range 30 days to 4.5 years post-HCT) (Table 1), and 4 went on to develop tMN 0.3 to 5.5 years post-HCT. In all cases, these patients had a normal karyotype prior to HCT but later presented with tMN characterized by *TP53* mutations and a complex karyotype. In contrast, tMN was not observed in any patient with successful engraftment.

**Table 1.** Patients receiving allogeneic transplantation for sickle cell disease.

Of the 4 patients that developed tMN, two patients (patients 1 and 3) were previously reported as *TP53* positive in the PB prior to HCT at a VAF of 0.34% and 0.06%, respectively^6^ (Table 1). Pre-HCT *TP53* NGS assay screening identified a pathogenic *TP53* variant (Arg273His) at a VAF of 0.36% in the blood of patient 4, corresponding to the mutation identified in the tMN presenting 0.3 years after HCT (Table 1, Supplementary Table 2).

We next examined the utility of *de novo TP53* mutation screening post-HCT in the patients that developed tMN using serial post-HCT PB or BM samples, where available (Table 1, Supplementary Table 2). For patient 1, the Arg175His *TP53* variant could be detected by mutation-specific ddPCR of PB collected 60 days post-HCT at a VAF of 0.035% and discovered by *TP53* NGS assay screening of BM collected 100 days post-HCT at a VAF of 0.75%, approximately 2 years prior to tMN development. For patient 2, who developed tMN 5.5 years after HCT but did not have a *TP53* mutation detected prior to transplant, the His179Arg variant could be detected by *TP53* NGS assay screening of blood at least six months prior to tMN development.

Interestingly, unlike pre-HCT samples, all post-HCT samples had additional pathogenic *TP53* variants detected beyond the variant ultimately responsible for tMN development (Supplementary Table 2). Notably, patient 2 had 4 post-HCT, pre-tMN samples available for screening, with multiple *TP53* mutations observed at each timepoint (VAF ranging from 0.03 to 0.99%), some which increase in VAF over the course of 5 years (Supplementary Figure 1). This pattern could be a generalizable phenomenon of prior chemotherapy/radiation exposure from HCT conditioning or might represent a mutational signature predictive of tMN development risk. Additional studies will be required to establish the clinical significance.

Importantly, in contrast, *TP53* NGS assay screening of PB DNA collected pre-HCT (range 18 days to 1 year) from the 15 patients who did not develop a tMN post-HCT, failed to identify any pre-existing pathogenic *TP53* variants (Table 1), indicating a low incidence of *TP53* mutations in this patient population prior to HCT.

Finally, while the *TP53* NGS screening assay used in this study was able to identify *TP53* mutations *de novo* down to approximately 0.1% VAF using a minimal sequencing read budget (∼10 million reads/sample), the variant discovery limit of this assay remains variable throughout the *TP53* gene due to technical limitations of single-stranded UMI error correction (range for average discovery limit at each hotspot position: 0.037% - 0.13%) (Figure 1D). Duplex sequencing, which utilizes double-stranded consensus sequences, can further reduce false-positive errors, allowing for an increased detection limit for *TP53* mutational screening. We tested the performance of duplex sequencing on 7 normal controls and found that when using the same DNA input as the single-stranded UMI method, the median detection limit was 0.012% (range 0.009-0.018%) across the *TP53* hotspot regions (Supplementary Figure 2). This nearly 5-fold increase in sensitivity along with improved consistency across the gene region could prove valuable for the future studies needed to establish the clinical significance of *TP53* mutational screening in patients at risk of developing tMN.

In summary, we report that NGS-based prospective screening for the reliable *de novo* discovery of low-level somatic *TP53* mutations. We successfully implemented this method to screen 19 patients with SCD who received HCT and found that no evidence of *TP53* mutations in patients who did not develop tMN. In contrast, patients who later developed tMN had pathogenic *TP53* mutations detectable both before and after HCT. This study provides generalizable evidence that ultra-deep NGS-based discovery of *TP53* mutations could be a valuable tool prior to treatments associated with a risk of subsequent tMN. Large scale prospective screening of patients at risk of tMN will now be necessary to validate the clinical utility of such a screening approach.

## METHODS

### Patient cohort

A total of 19 patients with SCD who underwent non-myeloablative peripheral blood HCT from June 2010 through October 2020 on protocols approved by the National Heart, Lung, and Blood Institute Institutional Review Board (Clinical Trials.gov identifiers NCT00061568, NCT02105766, NCT00977691, or NCT03077542) were included in this study. All subjects gave written informed consent. Details regarding the patient’s clinical characteristics, hematologic malignancy status, and clinical course were obtained by reviewing their medical records. For patients who developed a myeloid malignancy, somatic *TP53* mutations were originally identified by clinical next-generation sequencing panels at the time of myeloid malignancy diagnosis and confirmed by digital droplet PCR (ddPCR).

### Samples

DNA from peripheral blood (PB) was collected for all 19 SCD patients prior to HCT (range: 17 days to 1 year). For the 4 patients who developed a myeloid malignancy after HCT, DNA was collected from PB and/or bone marrow (BM) at various timepoints after HCT and at the time of myeloid malignancy diagnosis. For assay validation, genomic DNA (gDNA) from 3 positive control samples known to contain 7 unique *TP53* variants were diluted in a gDNA from healthy donor PB to achieve an anticipated variant allele fraction (VAF) ranging from 0.003% to 1%. The dilutions were prepared considering the variant with highest reported VAF for each sample.

Additionally, control DNA was used for establishing assay background using DNA extracted from healthy donor PB or Genome in a Bottle reference DNA (NA12878, NA24385, NA24149, NA24143, and NA24149) from Coriell Institute.

### Ultra-deep *TP53* DNA sequencing

A custom anchored multiplex PCR-based error-corrected targeted DNA sequencing panel (VariantPlex, ArcherDx, Boulder, CO) was designed with full coverage of coding regions of *TP53* gene. Libraries were prepared according to manufacturer’s instructions, with slight modifications. In short, 500-ng of gDNA was subjected to DNA fragmentation, end repair, A-tailing, purification using SPRIselect reagent (cat# B23318, Beckman Coulter, Inc., Brea, CA), and ligation with a universal ArcherDx molecular barcode (MBC) adapter, which tags each DNA molecule with a unique molecular index (UMI) and allows for unidirectional amplification of the sample using gene-specific primers. Following molecular barcode ligation, the libraries were subjected to two rounds of nested PCR for target enrichment. For the first PCR, amplification was performed as follows: 95°C for 3 minutes; 16 cycles of 95°C for 30 seconds, 65°C for 5 minutes; 72°C for 3 minutes. For the second PCR, amplification was performed as follows: 95°C for 3 minutes; 22 cycles of 95°C for 30 seconds, 65°C for 5 minutes; 72°C for 3 minutes.

The resulting next generation sequencing (NGS) libraries were subjected to paired-end 150-bp sequencing on a NovaSeq 6000 (Illumina, San Diego, CA), according to manufacturer’s instructions. Libraries were pooled for sequencing such that each sample had a unique dual index.

NGS libraries were prepared on the serial dilution samples, pre-HCT SCD patient PB, and post-HCT SCD patient PB/BM according to the manufacturer’s instructions. For the normal dataset, 6 NGS libraries were prepared using Genome in a Bottle reference DNA (NA12878, NA24385, NA24149, NA24143, and NA24149) from Coriell Institute.

### *TP53* NGS data analysis

Raw sequencing fastq files were analyzed using the ArcherDx Analysis software version 6.2.7 using the SNP-Indel pipeline as previously described^14^. Variants were called using both the *de novo* (LoFreq and Freebayes) and targeted (Vision) variant algorithms (using a list of common hotspot *TP53* mutations to improve detection).

Utilizing the normal dataset, the analysis software generates a background noise profile for each variant at each nucleotide position. The limit of detection (LOD) is expressed as the minimum allele fraction at which a variant can be distinguished from the underlying noise at a statistical power of 0.95, referred to as the 95 Minimum Detectable Allele Fraction (95 MDAF).

*TP53* variants underwent a first round of filtering to remove artifacts resulting from library preparation and sequencing as follows: deep alternate observations (DAO) ≥ 3, HRUN (maximum homopolymer length the variant resides in according to the reference sequence and alt sequence) ≤ 8, median distance of the variant to the start site generated by random ligation of the molecular barcode adapter ≤ 20, no sample strand or sequence direction bias, outlier *p* value for non- and error corrected (deep) allele fraction against the normal dataset: ≤ 10^-4^ and ≤ 10^-2^ respectively, and a reported population frequency in gnomAD database ≤ 0.001. Remaining variants with a predicted potentially deleterious variant consequence (i.e. missense, frameshift, splicing, indel) underwent manual curation to select for pathogenic or likely pathogenic mutations.

### Droplet digital PCR (ddPCR)

Assays were designed for the select *TP53* variants (TaqMan SNP Genotyping Assays, Thermo Fisher Scientific or ddPCR Mutation Detection Assay, Bio-Rad) and used to analyze 500ng of DNA from serial dilution samples and the DNA isolated from PB or BM of SCD patient samples using the QIAcuity One 5plex (Qiagen) or QX200 or QX600 Droplet reader (Bio-Rad) system. The data was analyzed using QIAcuity Software Suite or QuantaSoft Analysis Pro (1.0.596), QX Manager (Standard edition, 2.0).

### Duplex sequencing

*TP53* mutational profiling was also evaluated using targeted duplex sequencing, a DNA sequencing method that generates double-stranded duplex consensus sequencing. Targeted DNA sequencing was performed on 500-ng of healthy donor PB DNA utilizing the TwinStrand Duplex Sequencing AML-XP panel (TwinStrand Biosciences). Briefly, gDNA was enzymatically fragmented, followed by end repair, A-tailing, and DuplexSeq adapter ligation prior to amplification. Following indexing PCR, libraries were hybridized with biotinylated 120-mer DNA probes and purified with streptavidin magnetic beads. Following washes, additional PCR was performed, followed by another round of hybridization, capture, washes, and final PCR. Libraries were sequenced using paired-end 150bp sequencing on a NovaSeq 6000 (Illumina, San Diego, CA), according to manufacturer’s instructions. Alignment, duplex consensus sequence generation, and variant calling were performed utilizing the Twinstrand DuplexSeq AML XP workflow on the DNAnexus platform. A minimum of 2 duplex consensus reads were required for making a positive call and, in the absence of any duplex reads in healthy donor blood in hotspot regions the limit of detection at each position was defined as 2 divided by the duplex depth.

### Data visualization

NGS data and variants were visualized using R (version 4.3.1) in R Studio (version 2023.06.2 Build 561) and publicly available libraries on GitHub: ggplot2, dplyr, tidyverse, and trackViewer.

### Data availability

Raw sequencing data are available in the NCBI Sequence Read Archive (SRA) (Accession: XXX).

## Data Availability

All data produced in the present study are available upon reasonable request to the authors.

https://ashpublications.org/blood/article/135/14/1185/452511/Baseline-TP53-mutations-in-adults-with-SCD

## Acknowledgement

This work was funded by the Intramural Research Program of the National Heart, Lung, and Blood Institute of the National Institutes of Health.

