## Supplementary Figure 1 for "*TP53* mutation screening for patients at risk of myeloid malignancy"

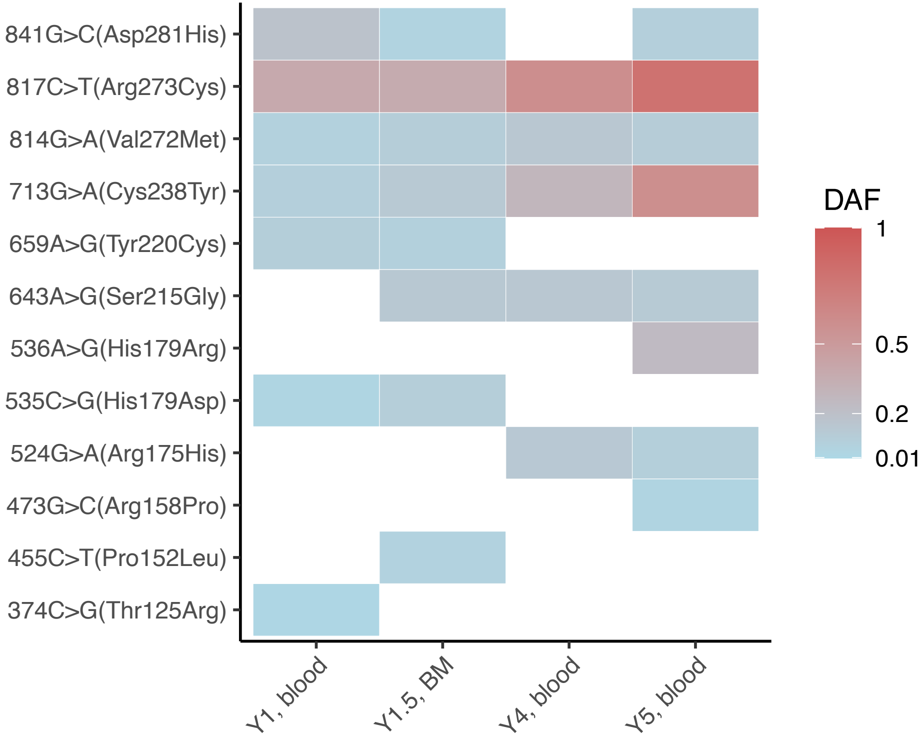


Supplementary Figure 1. Variant allele frequency (VAF) kinetics for the detected variants in patient 2.
