## Supplementary Figure 2 for "*TP53* mutation screening for patients at risk of myeloid malignancy"

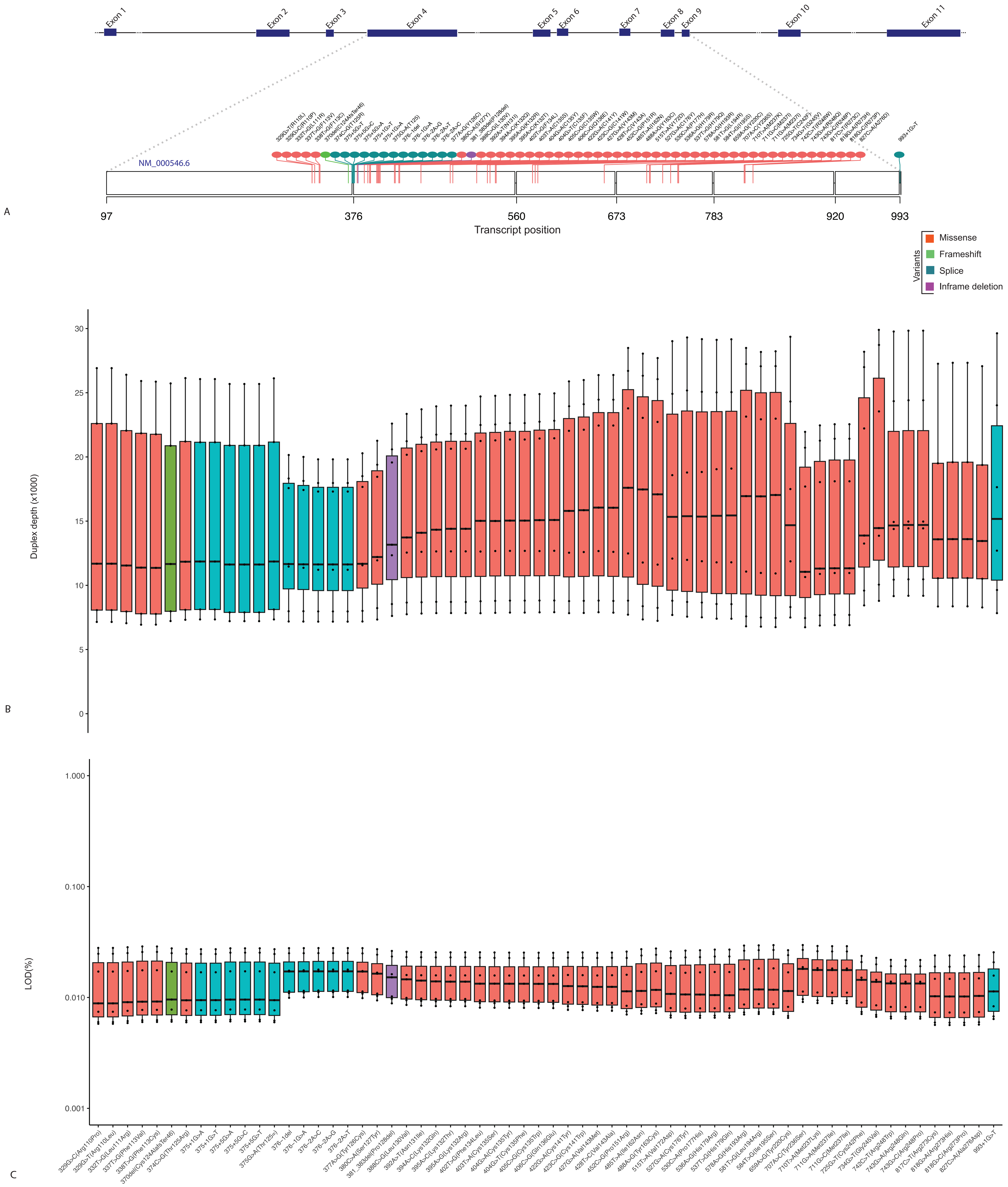


Supplementary Figure 2. A, Position of the tMN and AML hotspot across the TP53 gene and transcript. B, Depth and C, Limit of detection (LOD) of the duplex sequencing method.
